# Estimation of SARS-CoV-2 mortality during the early stages of an epidemic: a modeling study in Hubei, China, and six regions in Europe

**DOI:** 10.1101/2020.03.04.20031104

**Authors:** Anthony Hauser, Michel J. Counotte, Charles C. Margossian, Garyfallos Konstantinoudis, Nicola Low, Christian L. Althaus, Julien Riou

**Affiliations:** Institute of Social and Preventive Medicine, University of Bern, Bern, Switzerland; Department of Statistics, Columbia University, New York, NY; MRC Centre for Environment and Health, Department of Epidemiology and Biostatistics, School of Public Health, Imperial College London, London, UK; Division of infectious diseases, Federal Office of Public Health, Bern, Switzerland

## Abstract

**Background:** As of 16 May 2020, more than 4.5 million cases and more than 300,000 deaths from disease caused by severe acute respiratory syndrome coronavirus 2 (SARS-CoV-2) have been reported. Reliable estimates of mortality from SARS-CoV-2 infection are essential to understand clinical prognosis, plan health care capacity and for epidemic forecasting. The case fatality ratio (CFR), calculated from total numbers of reported cases and reported deaths, is the most commonly reported metric, but can be a misleading measure of overall mortality. The objectives of this study were to: 1) simulate the transmission dynamics of SARS-CoV-2 using publicly available surveillance data; 2) infer estimates of SARS-CoV-2 mortality adjusted for biases and examine the CFR, the symptomatic case fatality ratio (sCFR) and the infection fatality ratio (IFR) in different geographic locations.

**Method and Findings:** We developed an age-stratified susceptible-exposed-infected-removed (SEIR) compartmental model describing the dynamics of transmission and mortality during the SARS-CoV-2 epidemic. Our model accounts for two biases: preferential ascertainment of severe cases and right-censoring of mortality. We fitted the transmission model to surveillance data from Hubei province, China and applied the same model to six regions in Europe: Austria, Bavaria (Germany), Baden-Württemberg (Germany), Lombardy (Italy), Spain and Switzerland. In Hubei, the baseline estimates were: CFR 2.4% (95% credible interval [CrI]: 2.1-2.8%), sCFR 3.7% (3.2-4.2%) and IFR 2.9% (2.4-3.5%). Estimated measures of mortality changed over time. Across the six locations in Europe estimates of CFR varied widely. Estimates of sCFR and IFR, adjusted for bias, were more similar to each other but still showed some degree of heterogeneity. Estimates of IFR ranged from 0.5% (95% CrI 0.4-0.6%) in Switzerland to 1.4% (1.1-1.6%) in Lombardy, Italy. In all locations, mortality increased with age. Among 80+ year olds, estimates of the IFR suggest that the proportion of all those infected with SARS-CoV-2 who will die ranges from 20% (95% CrI: 16-26%) in Switzerland to 34% (95% CrI: 28-40%) in Spain. A limitation of the model is that count data by date of onset are required and these are not available in all countries.

**Conclusions:** We propose a comprehensive solution to the estimation of SARS-Cov-2 mortality from surveillance data during outbreaks. The CFR is not a good predictor of overall mortality from SARS-CoV-2 and should not be used for evaluation of policy or comparison across settings. Geographic differences in IFR suggest that a single IFR should not be applied to all settings to estimate the total size of the SARS-CoV-2 epidemic in different countries. The sCFR and IFR, adjusted for right-censoring and preferential ascertainment of severe cases, are measures that can be used to improve and monitor clinical and public health strategies to reduce the deaths from SARS-CoV-2 infection.

**Author summary:** *Why was this study done?:*

- Reliable estimates of measures of mortality from severe acute respiratory syndrome coronavirus 2 (SARS-CoV-2) infection are needed to understand clinical prognosis, plan health care capacity and for epidemic forecasting.
- The case fatality ratio (CFR), the number of reported deaths divided by the number of reported cases at a specific time point, is the most commonly used metric, but is a biased measure of mortality from SARS-CoV-2 infection.
- The symptomatic case fatality ratio (sCFR) and overall infection fatality ratio (IFR) are alternative measures of mortality with clinical and public health relevance, which should be investigated further in different geographic locations.

*What did the researchers do and find?:*

- We developed a mathematical model that describes infection transmission and death during a SARS-CoV-2 epidemic. The model takes into account the delay between infection and death and preferential ascertainment of disease in people with severe symptoms, both of which affect the assessment of mortality.
- We applied the model to data from Hubei province in China, which was the first place affected by SARS-CoV-2, and to six locations in Europe: Austria, Bavaria (Germany), Baden-Württemberg (Germany), Lombardy (Italy), Spain and Switzerland, to estimate the CFR, the sCFR and the IFR.
- Estimates of sCFR and IFR, adjusted for bias, were similar to each other and varied less geographically than the CFR. IFR was lowest in Switzerland (0.5%) and highest in Hubei province (2.9%). The IFR increased with age; among 80+ year olds, estimates ranged from 20% in Switzerland to 34% in Spain.

*What do these findings mean?:*

- The CFR does not predict overall mortality from SARS-CoV-2 infection well and should not be used for the evaluation of policy or for making comparisons between geographic locations.
- There are geographic differences in the IFR of SARS-CoV-2, which could result from differences in factors including emergency preparedness and response, and health service capacity.
- SARS-CoV-2 infection results in substantial mortality. Further studies should investigate ways to reduce death from SARS-CoV-2 in older people and to understand the causes of the differences between countries.

## Introduction

The pandemic of severe acute respiratory syndrome coronavirus 2 (SARS-CoV-2) infection has resulted in more than 4.5 million confirmed cases and more than 300,000 deaths from coronavirus disease 2019 (COVID-19), as of 16 May, 2020 [1]. The infection emerged in late 2019 as a cluster of cases of pneumonia of unknown origin in Wuhan, Hubei province [2, 3]. China had reported 84,038 cases and 4,637 deaths by 16 May, 2020, with no new deaths since early April. The largest outbreaks are now in the United States of America and Western Europe. The transmission characteristics of SARS-CoV-2 appear to be similar to those of the 1918 pandemic influenza strain [4], but, at this early stage of the pandemic, the full spectrum and distribution of disease severity and of mortality are uncertain. Reliable estimates of measures of mortality are needed to understand clinical prognosis, plan health care capacity and for epidemic forecasting.

The case fatality ratio (CFR), the number of reported deaths divided by the number of reported cases at a specific time point, is the most commonly used metric because most countries collect this information [2, 5]. However, the CFR can be misleading if used to assess the overall risk of death from an infection because of two opposing biases [6, 7]. First, because of the delay of several weeks between symptom onset and death, the number of confirmed and reported deaths at a certain time point does not consider the total number of deaths that will occur among already infected individuals (right-censoring). Second, surveillance-based case reports underestimate the total number of SARS-CoV-2-infected patients, because testing focuses on individuals with symptoms of COVID-19 and, among symptomatic cases, on patients with more severe manifestations (preferential ascertainment). In addition, the World Health Organization does not distinguish between SARS-CoV-2 infection and COVID-19 and defines a confirmed case as a person with laboratory confirmation of infection, irrespective of signs and symptoms. The number of cases detected and reported therefore depends on the extent and strategy of testing for SARS-CoV-2, especially amongst people without severe symptoms. Precisely defined measures could be more useful for describing SARS-CoV-2 mortality than the CFR [6]. The symptomatic case fatality ratio (sCFR) is the proportion of infected individuals showing symptoms who die over the course of their SARS-CoV-2 infection and is clinically relevant to assessment of prognosis and healthcare requirements. The infection fatality ratio (IFR) is the proportion of all people with SARS-CoV-2 infection who will eventually die from the disease, and is a central indicator for public health evaluation of the overall impact of an epidemic in a given population.

Estimates of the sCFR and IFR can be obtained from prospective longitudinal studies of representative samples of individuals with SARS-CoV-2 infection but such studies cannot provide the information needed for clinical and public health decision-making in real time. The objectives of this study were to: 1) simulate the dynamics of transmission and mortality of SARS-CoV-2 using publicly available surveillance data; and 2) provide overall and age-stratified estimates of sCFR and IFR for SARS-CoV-2, adjusted for right-censoring and preferential ascertainment, in different geographic locations.

## Methods

We developed an age-stratified susceptible-exposed-infected-removed (SEIR) compartmental model that describes the dynamics of SARS-CoV-2 transmission and mortality. We fitted the model to surveillance data from Hubei province (China) and six geographic locations locations in Europe: Austria, Bavaria (Germany), Baden-Württemberg (Germany), Lombardy (Italy), Spain and Switzerland. There is no written prospective protocol for the study. The analysis has been developed specifically for the research question, and adapted in response to peer review comments. Main changes include assuming pre- and asymptomatic transmissions and running additional sensitivity analyses. In the revised version, the analysis also includes additional regions, for which data had been made available. All code including the different versions of the model and manuscript are available from https://github.com/jriou/covid_adjusted_cfr. This study is reported as per the TRIPOD guideline (S2 Text).

### Setting and data, Hubei province, China

The first known case of SARS-CoV-2 infection was traced back to December 1st, 2019 in Wuhan, the main city of Hubei province, China [3]. The first reported death was on 11 January 2020. Human-to-human transmission of SARS-CoV-2 led to exponential growth of the reported incidence of cases (Fig 1A). On 20 January 2020, Chinese authorities implemented extensive control measures in Hubei: early identification and isolation of clinical cases, tracing and quarantining of contacts, temperature checks before accessing public areas, extension of the lunar new year holiday period, and extreme social distancing, including cancellation of mass gatherings [8]. Three days later, a cordon sanitaire was imposed, with strict traffic restrictions. From 27 January, the daily incidence of cases, by date of symptom onset, started to plateau, then decreased.

**Fig 1.**
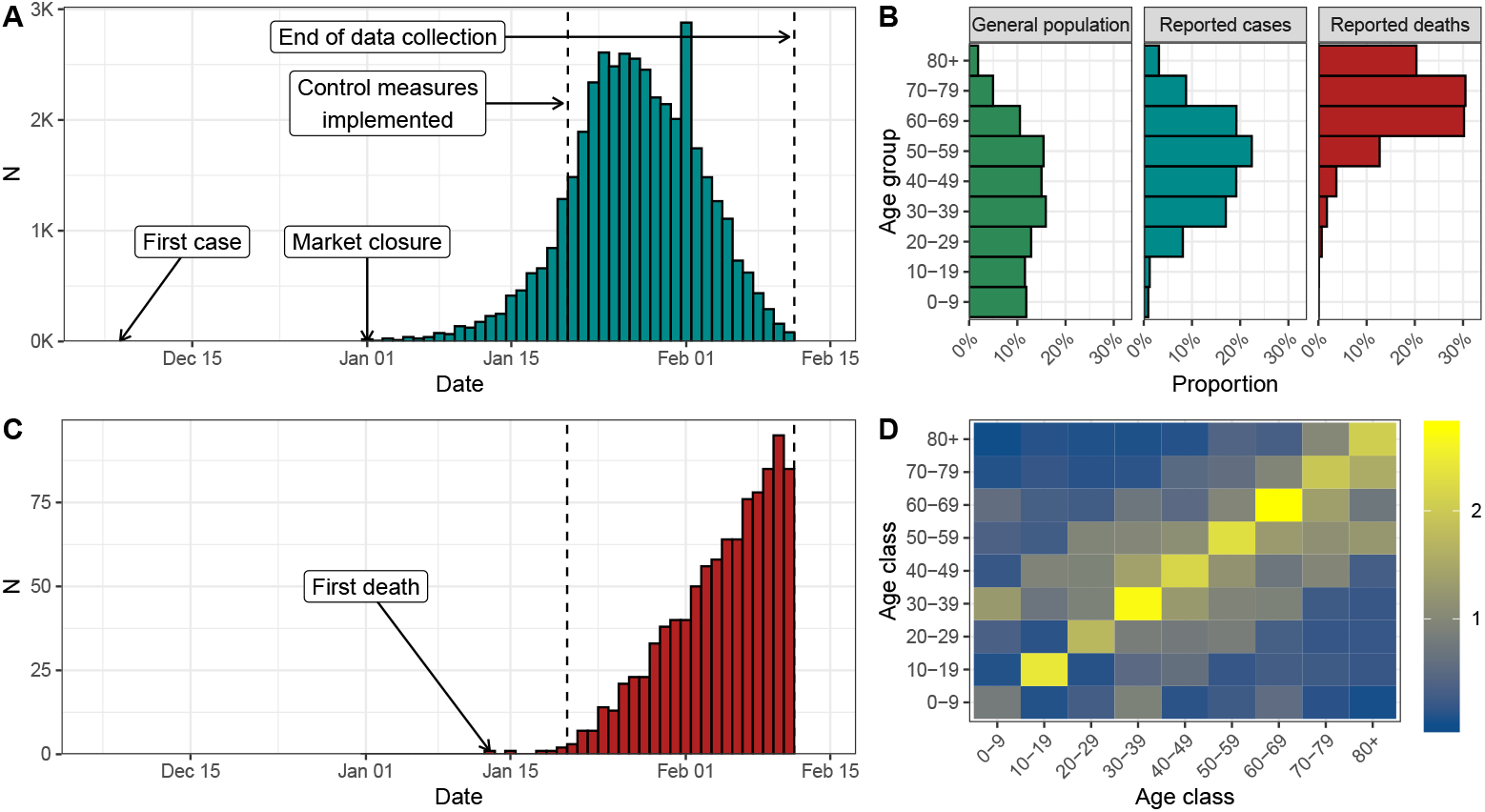
(A) Reported number of confirmed cases of SARS-CoV-2 infection by date of symptom onset in Hubei, China until 11 February 2020. (B) Age distribution of the Chinese population and of the reported cases and deaths in Hubei, China. (C) Reported number of deaths associated with SARS-CoV-2 infection in in Hubei, China until 11 February 2020. (D) Age-specific contact matrix from a 2018 survey conducted in Shanghai, China [11] applied to Hubei province.

The Chinese Center for Disease Control and Prevention (China CDC) reported the number of cases by date of symptom onset, and the age distribution of cases and deaths up 11 February 2020 in China (Fig 1B) [9]. We extracted these data, together with the age distribution of the Chinese population. Deaths counts were obtained from a repository aggregating data from Chinese public data sources [10]. We used data about the daily number of potentially infectious contacts by age group in Shanghai [11]. We assumed that all data sources were applicable to the population of Hubei. As of 11 February, after which information about date of symptom onset was no longer available, there were 41,092 cases and 979 deaths, resulting in a CFR of 2.4%.

### Setting and data, six geographic locations in Europe

The first cases of SARS-CoV-2 infection in Europe were reported at the end of January 2020. Italy was the first country with a large epidemic, after a cluster of cases, followed shortly by the first deaths, emerged in Lombardy at the end of February. As of 16 May 2020, Europe is the continent having reported the highest number of cases and deaths (more than 1,800,000 confirmed cases and 160,000 deaths) [1].

We selected European countries that reported the daily number of cases of confirmed SARS-CoV-2 infection by date of symptoms onset. In countries where this information was available at a regional level, we selected worst-affected regions. We extracted data about the number of confirmed cases by symptom onset, the daily number of deaths, and the distribution of cases and deaths across age groups for each of six locations: Austria, Bavaria (Germany), Baden-Württemberg (Germany), Lombardy (Italy), Spain and Switzerland.

For Austria, we obtained all required data from published reports from 11 March to 14 April [12]. On 14 April, there were 14,151 reported cases and 399 deaths (CFR 2.8%). For Germany, we used published data from 3 March to 16 April [13]. Age distributions, available at the country level only, were applied to both regions. On 16 April, there were 31,196 cases (62% with date of onset) and 802 deaths in Baden-Württemberg (CFR 2.6%) and 36,538 cases (56% with date of onset) and 1,049 deaths in Bavaria (CFR 2.9%). For Lombardy, we collected published data from 11 February to 25 April [14, 15]. Age distributions at the national level were applied to Lombardy. On 25 April, there were 74,346 cases (77% with date of onset) and 13,263 deaths (CFR 17.8%). For Spain, we used published data from 2 March to 16 April [16]. On 16 April, 178,031 cases (79% with date of onset) and 19,478 deaths were reported (CFR 10.9%). For Switzerland, we used individual-level data from 2 March to 23 April aggregated by day of onset or day of death [17]. On 23 April, there were 33,228 cases (11% with date of onset) and 1,302 deaths (CFR 3.9%). Further details about the data are available in S1 Text, section 1.

### Age-structured model of SARS-CoV-2 transmission and mortality

We used an age-stratified susceptible-exposed-infected-removed (SEIR) compartmental model that distinguished between incubating, pre-symptomatic, asymptomatic and symptomatic infections. We stratified the population into nine 10-year groups (0-9 up to 80+ years) for all locations except Austria where the age groups were 0-4, 5-14, up to 75+ years. We assumed that susceptibility to SARS-CoV-2 and the risk of acquisition per contact is identical for each age group and that transmission is possible during pre-symptomatic and asymptomatic infections. We used age-specific contact matrices to model contact patterns according to age group (contact matrix derived by Zhang et al. for Hubei [11], and the POLYMOD contact matrix for the six European locations [18]). We modelled the decrease in SARS-CoV-2 transmission due to control measures using a logistic function for the transmission rate.

In the model, after an average incubation period of 5.0 days [19], 81% (95%CrI: 71-89) of infected people develop symptoms of any severity and the rest remain asymptomatic [20, 21]. The estimated proportion of symptomatic infections was derived from outbreak investigations included in a systematic review and is implemented as a beta distribution to propagate uncertainty. Studies that have estimated the proportion of asymptomatics have not provided conclusive evidence of an age trend, so we assumed it to be constant [22]. We assumed reduced infectiousness during the period of 2.3 days preceding symptom onset (pre-symptomatic compartment) and also among asymptomatic individuals [19].

The model was used to compute the number of symptomatic SARS-CoV-2 infections by day of symptom onset in each age group. We applied an age-specific ascertainment proportion to the number of symptomatic infections to estimate the number of reported cases of SARS-CoV-2 infections by date of symptom onset. To identify the parameters, we assumed that 100% of infections in the oldest age group (80+ years old, 75+ years old in Austria) were reported. We assumed that mortality only occurred in symptomatic people, and that the time from symptom onset to death followed a log-normal distribution with mean 20.2 days and standard deviation 11.6 [23]. This allowed us to account for the deaths occurring after the date of data collection.

Separately for Hubei and the six European locations, we simultaneously fitted our model to the data sets described above (Fig 1): (1) the number of confirmed cases by day of symptom onset, (2) the number of deaths by day of occurrence, (3) the age distribution of all confirmed cases and (4) the age distribution of all reported deaths. We assumed a negative binomial distribution for data (1) and (2), and a multinomial distribution for data (3) and (4). All parameters were estimated from data except for the incubation period, the generation time, the contribution of presymptomatics to transmission, the presymptomatic duration, and the time from symptom onset to death.

The fitted model was used to produce estimates (median posterior distributions with 95% credible intervals, CrI) of the total number of symptomatic and pre-/a-symptomatic infections (adjusted for preferential ascertainment) and of the total number of deaths (adjusted for right-censoring). These were then transformed into adjusted estimates of sCFR and IFR. Besides parameter values and model structure, these estimates rely on the following additional assumptions:

1. The severity of symptoms differs by age group and influences the probability of reporting;
2. All deaths due to SARS-CoV-2 infection have been identified and reported;
3. The susceptibility to SARS-CoV-2 infection is identical across age groups;
4. The average standard of care is stable for the period of interest and the next two months, during which a proportion of the infected people will eventually die;
5. The ascertainment probability by age is constant over the periods considered. Further details about the method are available in S1 Text, section 2.

### Sensitivity analysis

From 12 February 2020, the Chinese authorities changed their criteria for reporting cases, increasing the total by more than 25,000. Reported numbers of deaths increased on 16 April, when Wuhan city reported an additional 1,290 deaths. We ran a sensitivity analysis with corrected numbers of cases and deaths in Hubei province. We also examined the impact of assuming a 50% lower susceptibility in 0-19 years old, and of a lower ascertainment among 80+ year olds (from 90% to 10%, compared with a fixed proportion of 100% in the main analysis). We also re-fitted the model at different dates of data collection (every 5 days from 12 January to 11 February) to examine the effect of the accumulation of data over time. Additional sensitivity analyses are presented in S1 Text, section 6.

We implemented the model in a Bayesian framework using Stan [24]. All code and data are available from https://github.com/jriou/covid_adjusted_cfr.

## Results

Our model accurately describes the dynamics of transmission and mortality by age group during the SARS-CoV-2 epidemic in Hubei from 1 January to 11 February 2020 (Fig 2). The model predicts that control measures implemented from 20 January reduced SARS-CoV-2 transmissibility by 92% (95%CrI: 87-100), with a steep diminution in case incidence 4.3 (95%CrI: 3.2-5.4) days after 20 January. Assuming 100% of cases aged 80 and older were initially reported, we estimate that a total of 83,300 individuals (95%CrI: 73,000-98,600) were infected in Hubei between 1 January and 11 February 2020. Of these, the number of symptomatic cases is estimated at 67,000 (95%CrI: 60,500-73,600), 1.6 times (95%CrI: 1.5-1.8) more than the 41,092 reported cases during that period. Accounting for the later correction in the number of reported cases, the total number of infections increases to 138,000 (95%CrI: 120,000-162,000). The proportion of ascertained cases by age group increased from less than 9% (95%CrI: 8-10) under 20 years old to 93% (95%CrI: 88-98) in the age group 70-79 (the ascertainment proportion was assumed to be 100% in the age group 80+, Fig 3A).

**Fig 2.**
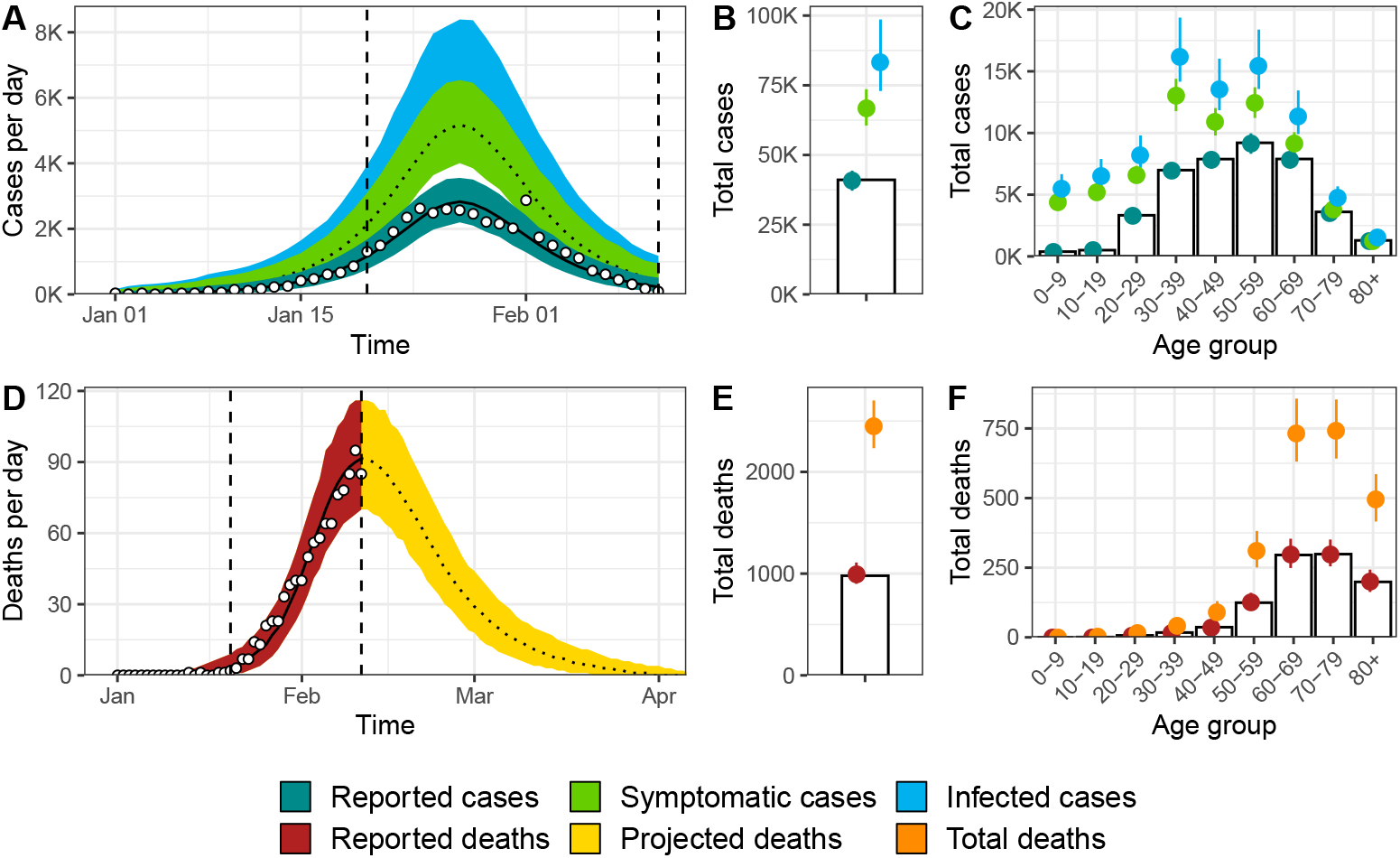
Model fit for Hubei, China of (A) incident cases of SARS-CoV-2 infection by date of symptom onset, (B) total cases, (C) age distribution of cases, (D) incidence of deaths, (E) total number of deaths among individuals infected until 11 February 2020 and (F) age distribution of deaths. White circles and bars represent data. Lines and shaded areas or points and ranges show the posterior median and 95% credible intervals for six types of model output: reported cases, symptomatic cases, overall cases (i.e. symptomatic and asymptomatic cases), reported deaths until 11 February 2020, projected deaths after 11 February 2020 and overall deaths.

**Fig 3.**
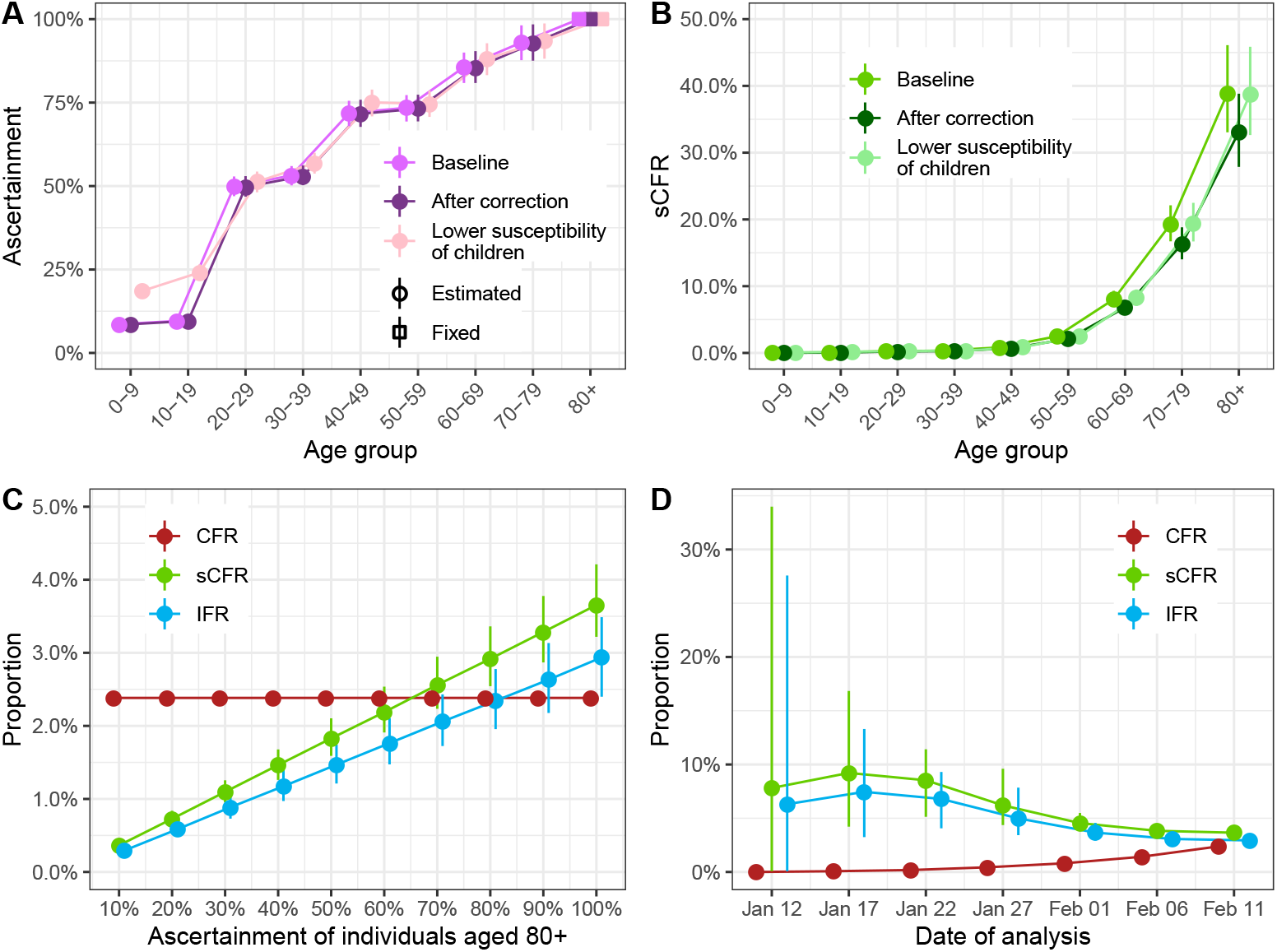
(A) Estimated proportion of cases ascertained by age group in Hubei, China (baseline, after the later correction of the number of reported cases and deaths, and assuming 50% lower susceptibility of children aged 0-19). (B) Estimated case symptomatic fatality ratio by age group in Hubei, China. (C) Impact of varying the fixed proportion of cases ascertained among individuals aged 80 and older from 10% to 100% on the mortality estimates. (D) Mortality estimates at different dates of reporting (every 5 days from January 12 to February 11).

The model predicts a total of 2,450 deaths (95%CrI: 2,230-2,700) among all people infected until 11 February in Hubei (compared with 979 deaths at this point without adjusting for right-censoring). This results in an estimated IFR of 2.9% (95%CrI: 2.4-3.5, Table 1). Assuming the later correction of deaths was evenly distributed by date of symptom onset and age group, the total number of deaths increases to 3,430 (95%CrI: 3,120-3,760). When using the corrected numbers of cases and deaths, we derived an IFR of 2.5% (95%CrI: 2.1-2.9).

**Table 1.**
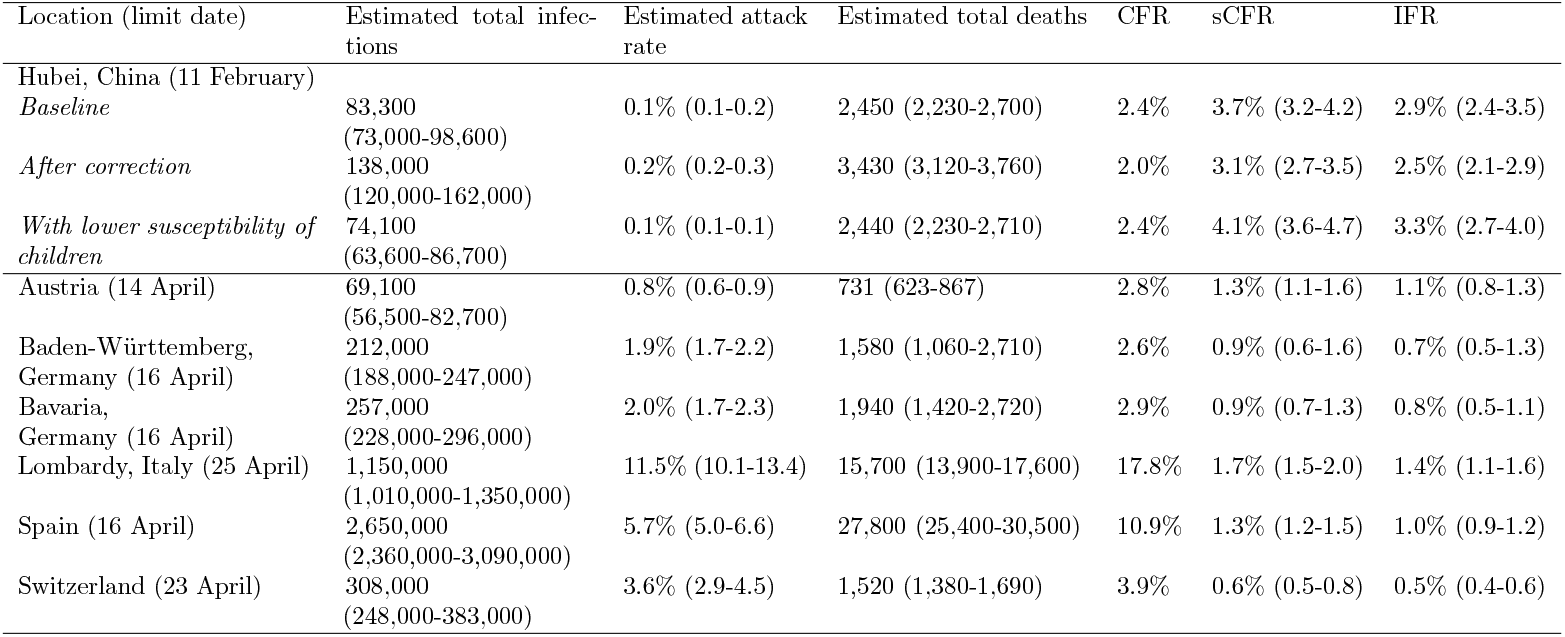
Model estimates of total infections of SARS-CoV-2 infection, attack rate, total deaths, case fatality ratio (CFR), symptomatic case fatality ratio (sCFR) and infection fatality ratio (IFR) by location until the limit date

| Location (limit date) | Estimated total infections | Estimated attack rate | Estimated total deaths | CFR | sCFR | IFR |
| --- | --- | --- | --- | --- | --- | --- |
| Hubei, China (11 February) |  |  |  |  |  |  |
| *Baseline* | 83,300<br>(73,000-98,600) | 0.1% (0.1-0.2) | 2,450 (2,230-2,700) | 2.4% | 3.7% (3.2-4.2) | 2.9% (2.4-3.5) |
| *After correction* | 138,000<br>(120,000-162,000) | 0.2% (0.2-0.3) | 3,430 (3,120-3,760) | 2.0% | 3.1% (2.7-3.5) | 2.5% (2.1-2.9) |
| *With lower susceptibility of children* | 74,100<br>(63,600-86,700) | 0.1% (0.1-0.1) | 2,440 (2,230-2,710) | 2.4% | 4.1% (3.6-4.7) | 3.3% (2.7-4.0) |
| Austria (14 April) | 69,100<br>(56,500-82,700) | 0.8% (0.6-0.9) | 731 (623-867) | 2.8% | 1.3% (1.1-1.6) | 1.1% (0.8-1.3) |
| Baden-Württemberg, Germany (16 April) | 212,000<br>(188,000-247,000) | 1.9% (1.7-2.2) | 1,580 (1,060-2,710) | 2.6% | 0.9% (0.6-1.6) | 0.7% (0.5-1.3) |
| Bavaria, Germany (16 April) | 257,000<br>(228,000-296,000) | 2.0% (1.7-2.3) | 1,940 (1,420-2,720) | 2.9% | 0.9% (0.7-1.3) | 0.8% (0.5-1.1) |
| Lombardy, Italy (25 April) | 1,150,000<br>(1,010,000-1,350,000) | 11.5% (10.1-13.4) | 15,700 (13,900-17,600) | 17.8% | 1.7% (1.5-2.0) | 1.4% (1.1-1.6) |
| Spain (16 April) | 2,650,000<br>(2,360,000-3,090,000) | 5.7% (5.0-6.6) | 27,800 (25,400-30,500) | 10.9% | 1.3% (1.2-1.5) | 1.0% (0.9-1.2) |
| Switzerland (23 April) | 308,000<br>(248,000-383,000) | 3.6% (2.9-4.5) | 1,520 (1,380-1,690) | 3.9% | 0.6% (0.5-0.8) | 0.5% (0.4-0.6) |

The estimated sCFR, more relevant to the clinical setting, was 3.7% (95%CrI: 3.2-4.2) in the baseline analysis and 3.1% (95%CrI: 2.7-3.6) after correction of the increase number of reported cases and deaths. The estimated sCFR increased with age (S1 Text, section 5): under 20 years of age, below 1 in 1,000; 20-49 years, between 3 and 8 per 1,000; 50-59 years, 2.5% (95%CrI: 2.0-3.0); 60-69 years, 8.0% (95%CrI: 6.9-9.3); 70-79 years, 19.3% (95%CrI: 16.7-22.1); 80 years and older, 39.0% (95%CrI: 33.1-46.1).

In sensitivity analyses, the correction of the number of reported cases (+65%) and deaths (+40%) by the local authorities in Hubei did not influence the ascertainment proportion (Fig 3A) but led to a proportional decrease of the sCFR and IFR estimates by 15%, as expected from the correction applied (1.40*/*1.65 = 0.85, Fig 3B and Table 1). Second, lowering the susceptibility of individuals aged 0-19 years by 50% did not affect the ascertainment proportion or the sCFR in other age groups (Fig 3A and B). The decrease in the denominator led to a proportional increase of total sCFR and IFR. Third, relaxing the assumption of complete reporting of cases among individuals aged 80 years and older resulted in a proportional decrease of the sCFR and IFR (Fig 3C). Fourth, patterns in the observed mortality changed as the epidemic progressed (Fig 3D). The CFR increased as delayed deaths were reported. The sensitivity analysis suggests that our proposed approach overestimates the sCFR and IFR when applied before the peak of incidence (around 27 January) and stabilizes afterwards. Additional sensitivity analyses examining how the contribution of pre-symptomatic transmission, the susceptibility of children and several other choices in model structure and parameter values did not impact the results (S1 Text, section 6).

We applied the same model to data from six European locations with all required data: Austria, Germany (Baden-Württemberg and Bavaria), Italy (Lombardy), Spain and Switzerland. The model fit was satisfactory in all cases (S1 Text, section 3). CFR estimates differed widely between countries, while sCFR and IFR estimates were more similar to each other (Table 1 and Fig 4A). Across countries, model estimates of IFR ranged from 0.5% (95%CrI: 0.4-0.6) in Switzerland to 1.4% (95%CrI: 1.1-1.6) in Lombardy, Italy. The patterns of age-specific IFR estimates were similar across locations (Fig 4B), despite differences in the surveillance-reported age distribution of cases (Fig 4C). Some degree of variability remained between age-specific IFR estimates, especially in older age groups. Compared with Hubei province, higher proportions of cases in European locations were in older age groups, suggesting higher levels of preferential ascertainment of severe cases. This appears in the estimated patterns of the age-specific ascertainment proportion, with a generally lower ascertainment of age groups 20-79 in Europe compared to Hubei (Fig 4D). Additional results are presented in S1 Text, section 5.

**Fig 4.**
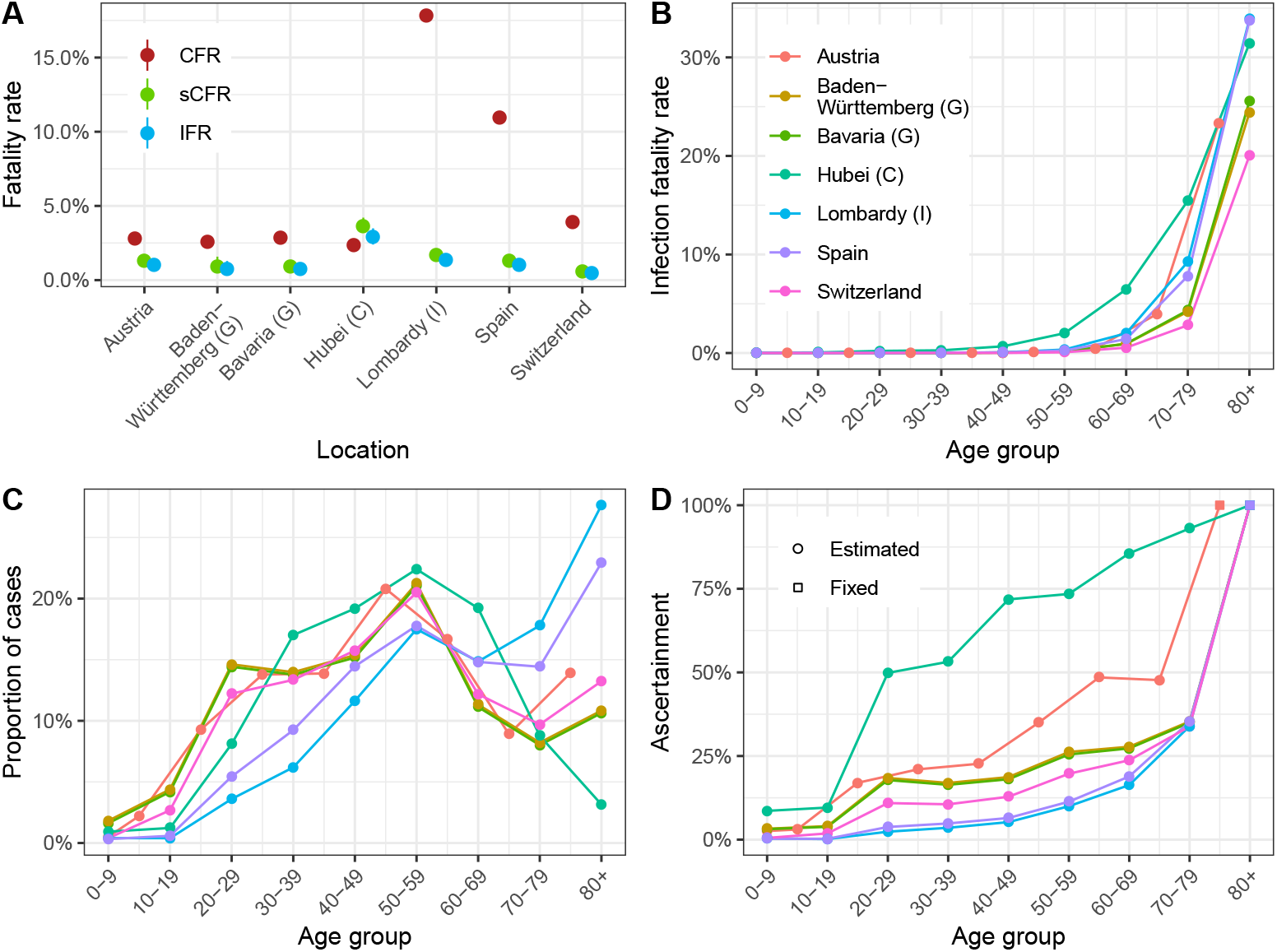
(A) Case fatality ratio, symptomatic case fatality ratio and infection fatality ratio estimates by geographic location. (B) Infection fatality ratio estimates by age group and location (for Austria, the estimates are adapted to the available age groups from 0-4 to 75+ years). (C) Proportion of cases ascertained by age group and location (color code as for panel B). (D) Distribution of reported cases by age group by location (color code as for panel B).

## Discussion

In this modelling study, we estimate different measures of mortality from SARS-CoV-2 infection in Hubei province, China and six geographic locations in Europe. After adjusting for right-censoring and preferential ascertainment, we estimate the IFR in Hubei to 2.9% (2.4-3.5), higher than the CFR of 2.4%. In different European settings, estimates of IFR ranged from 0.5% (95%CrI: 0.4-0.6) in Switzerland to 1.4% (95%CrI: 1.1-1.6) in Lombardy, compared with CFR of 3.9% and 17.8%, respectively. The model estimates of mortality show a strong age trend in all locations, with very high risks in people aged 80 years and older: between 20% (95%CrI: 16-26) in Switzerland and 34% (95%CrI: 28-40) in Lombardy.

### Strengths and limitations

Our work has four important strengths. First, we distinguish between the crude CFR and two separate measures of mortality, sCFR and IFR. Second, we use a mechanistic model for the transmission of SARS-CoV-2, and the mortality associated with SARS-CoV-2 infection which directly translates the data-generating mechanisms leading to biased observations of the number of deaths (because of right-censoring) and of cases (because of preferential ascertainment). Our model also accounts for the effect of control measures on disease transmission. We implemented the model in a Bayesian framework in order to propagate most sources of uncertainty from data and parameter values into the estimates. In Hubei, as the model captured most of the epidemic wave, the predicted number and timing of deaths could be compared with later reports of SARS-CoV-2 deaths, providing some degree of external validation (S1 Text, section 4). Third, our model is stratified by age group, which has been shown as a crucial feature for modelling emerging respiratory infections [25]. Fourth, the model uses surveillance data that can be collected routinely, and does not require individual-level data or studies in the general population.

Our study has several limitations. First, an important assumption is that all cases in symptomatic individuals aged 80 years and older were reported, as a result of more severe symptoms at older ages. We cannot confirm this, but the high risk of death from SARS-CoV-2 infection amongst the elderly was reported very early on [3], so we believe that most old people with symptoms sought care. Sensitivity analyses show that sCFR and IFR estimates decrease linearly with a lower ascertainment of infections among individuals aged 80 years and older (Fig 3C). For the IFR in Hubei province to be below 0.5%, fewer than 15% of infections in individuals aged 80 years and older would have been ascertained by the local authorities.

Second, our model requires surveillance data about the incidence of cases of reported SARS-CoV-2 by date of symptom onset. When information on symptom onset is only available for a subset of cases, we have to assume that data are missing at random. Additionally, within a given geographic location, the model assumes a constant ascertainment proportion and a constant mortality for each age group during the period. When repeating the analysis at different stages of the epidemic in Hubei, we found that estimates obtained before the epidemic peak led to overestimation of the sCFR and IFR (Fig 3D). This finding could be the result of a decrease in mortality as the epidemic progresses, but could also be attributed to a lower ability of the model to estimate the epidemic size before epidemic peak is reached, a common problem in epidemic modelling [26].

Third, we assume that the deficit of reported cases among younger age groups is a result of preferential ascertainment, whereby younger individuals have milder symptoms and are less likely to seek care, and does not reflect a lower risk of infection in younger individuals. During the pandemic of H1N1 influenza, lower circulation in older individuals was attributed to residual immunity [27]. There is no indication of pre-existing immunity to SARS-CoV-2 in humans [8]. Lower susceptibility of younger individuals for immunological reasons seems unlikely. Different contact patterns could contribute to different attack rates by age group. We include age-specific contact patterns in the model, so our results are dependent on the contact matrix used. In a sensitivity analysis, with 50% reduced susceptibility in children, the estimates of age-specific IFR in other age groups did not change, but the lower number of total infections and led to a higher IFR.

Fourth, the proportion of SARS-CoV-2 infections that remains asymptomatic is still uncertain. To propagate this uncertainty into the results, we implemented a prior distribution informed by the findings of outbreak investigations included in a living systematic review and meta-analysis [20, 21]. Our estimate is in agreement with the findings of a statistical modelling study of an outbreak on the cruise ship “Diamond Princess”, estimating an average proportion of symptomatic infections of 82.1% (95%CrI: 79.8-84.5) [28]. Another study of 87 contacts of infected cases in Shenzhen, China, estimated that 80.4% (95%CrI: 70.9-87.4) were symptomatic [19]. Additionally, dichotomization into asymptomatic and symptomatic is a simplification; SARS-CoV-2 causes a spectrum of symptoms, likely depending on age, sex and comorbidities. Serological surveys in the general population will be needed to better characterize asymptomatic infections [29].

### Comparison with other studies

Our model-based estimates have some degree of external validation from serological studies of previous exposure to SARS-CoV-2. In Geneva, Switzerland, a study reported an attack rate of 9.7% (95% confidence interval: 6.1-13.1) in the city, resulting in an IFR of 0.6%, very close to our national estimate for Switzerland [30]. Preliminary results from a national seroprevalence study in Spain, with more than 60,000 participants, found an attack rate of 5.0% (95% confidence interval: 4.7-5.4), consistent with our estimate of 5.7% (95% CrI: 5.0-6.6) [31]. A study of excess mortality in Italy estimated 17,786 *±* 269 deaths in Lombardy, close to our credibility interval for the number of total deaths [32]. This study did not attempt to estimate the size of the epidemic, but only applied the proportion of positive tests to the population to obtain an upper limit of epidemic size, which resulted in a lower bound for the IFR of 0.6% in Lombardy.

Model-based estimates of mortality from SARS-CoV-2 in China, adjusting for bias, vary. Our estimate for Hubei province is higher than the sCFR of 1.4% estimated in two other modelling studies [33, 34]. Differences in modelling approaches and assumptions explain the variation. Verity et al. used a similar modelling approach, but applied their findings to all of mainland China, where mortality outside Hubei province appeared lower [9]. This paper also assumed a homogeneous attack rate across age groups, rather than simulating epidemics using an age-specific contact matrix. Wu et al. used another approach, assuming that susceptibility to infection varies by age. Both Verity et al. and Wu et al. used data from individuals leaving Wuhan before lockdown was implemented to infer ascertainment, where we fixed it to 100% for the oldest age group. This resulted in comparatively lower ascertainment proportions (up to 70% for the oldest age groups in Verity et al., 2% for Wu et al.), and consequently to higher estimates of epidemic size and lower estimates of sCFR. The use of data from travellers might result in bias, especially if people who can travel are healthier than the general population.

Other studies that attempt to adjust for right-censoring of deaths give different estimates of mortality in China than in our study. A study using a competing risk model estimated mortality at 7.2% (95% confidence interval: 6.6%-8.0%) for Hubei province [35]. Using data on exported cases, another team estimated mortality of 5.3% (95% confidence interval: 3.5%, 7.5%) among confirmed cases in China [36]. Another team reported a CFR of 18% (95% credible interval: 11-81%) among cases detected in Hubei, accounting for the delay in mortality and estimated the IFR at 1.0% (95% CI: 0.5%-4%), based on data from the early epidemic in Hubei and from cases reported outside China [37]. Our estimate of mortality among all infected cases in Hubei is also higher than in an earlier version of this work (2.9% against 1.6%) [38]. We believe the newer estimate to be more reliable for two reasons. First, we implemented age-specific risks of transmission through a contact matrix, which partially explains the age patterns in reported SARS-CoV-2 infections and leads to lower estimates of the total number of infections, thus increasing mortality. Second, a higher estimated proportion of symptomatic people based on new data also led to higher estimates of mortality among all infected.

### Interpretation and implications

In this study, we propose a comprehensive solution to the estimation of mortality from surveillance data during outbreaks, using two measures of mortality [6]. Adjusted for right-censoring and preferential ascertainment of severe cases, the IFR is a measure of overall mortality associated with SARS-CoV-2 infection, which can be used to assess the potential consequences of the pandemic, e.g. using theoretical estimates of final epidemic size [39]. The sCFR is a measure of mortality that is most relevant to the clinical setting, for assessment of prognosis and prioritization of health care services. Crude CFR estimates are a poor predictor of mortality from SARS-CoV-2 infection, as demonstrates for instance the comparison of CFR and IFR values in Switzerland (high CFR, lowest IFR) and Hubei (lowest CFR, highest IFR). In addition to the inherent biases, the wide variation in CFR between geographic locations is likely to reflect external factors, including policies for testing and differences in systems of surveillance and reporting more than differences in mortality. Crude CFR values should therefore not be used for evaluating policy or making comparisons across settings.

Our model-based estimates of the IFR and sCFR varied geographically (Fig 4A). The highest estimate of IFR was found in Hubei province (2.9%; 95%CrI: 2.4-3.5 in the baseline analysis). The steep increase in mortality among people aged 60 years and older, reaching very high values in people aged 80 years and older is of concern. The credibility of this estimate, and of our approach for adjusting for right-censoring, is supported by the model predictions of reported daily SARS-CoV-2-associated deaths in Hubei province after 11 February (S1 Text, section 4). The estimated IFR decreases to 2.5% (95%CrI: 2.1-2.9) when accounting for the later correction of reported cases and deaths by the local authorities, and increases to 3.3% (95%CrI: 2.7-4.0) if we consider a lower susceptibility of individuals under 20 years. We also show that applying our model at earlier stages of the epidemic would have resulted in higher estimates of sCFR and IFR, and more uncertainty (Fig 3D). However, our estimates here correspond to an average value over the considered period, and it has been shown that mortality has changed over time as a result of an improvement of the standard of care [8].

The estimated IFR in Lombardy, 1.4% (95%CrI: 1.1-1.6), was lower than in Hubei province, but higher than in five other European locations. Further research is necessary to better understand the factors associated with SARS-CoV-2 mortality. These differences highlight the importance of local factors on the outcome of SARS-CoV-2 infection, including demographic characteristics. A partial explanation for the remaining heterogeneity is the lower degree of preparedness and health service capacity in northern Italy, which in Europe was affected first by the SARS-CoV-2 epidemic. Consequently, we suggest that a single mortality estimate should not be applied to all settings to estimate the total size of the epidemic [40]. This study shows that the IFR and sCFR, adjusted for right-censoring and preferential ascertainment biases, are appropriate measures of mortality for SARS-CoV-2 infection, which can be used to improve and monitor clinical and public health strategies to reduce the deaths from SARS-CoV-2 infection.

## Conclusions

We developed a mechanistic approach to correct the CFR for bias due to right-censoring and preferential ascertainment and provide adjusted estimates of mortality due to SARS-CoV-2 infection by age group. We applied this approach to seven different settings, showing that widely different estimates for the CFR corresponded in fact to more similar estimates of the IFR, around 3% in Hubei province, China, and ranging between 0.5 and 1.4% in six included European locations. Despite these similarities, substantial heterogeneity remains in the IFR estimates across settings, indicating the impact of local conditions on the outcome of SARS-CoV-2 infection. The steep increase in mortality among people aged 60 years and older, reaching very high values in people aged 80 years and older is of concern.

## Data Availability

All code and data are available from https://github.com/jriou/covid_adjusted_cfr

## Acknowledgments

We warmly thank Ben Bales for his help with the implementation of the model. We also thank the Chinese Center for Disease Control and Prevention, the Austrian Bundesministerium für Soziales, Gesundheit, Pflege und Konsumentenschutz, the German Robert Koch Institute, the Italian Dipartimento della Protezione Civile and Istituto Superiore di Sanita, the Spanish Ministerio de Sanidad and the Swiss Federal Office of Public Health for collecting the data and making it publicly available. Computations were conducted on UBELIX (http://www.id.unibe.ch/hpc), the high performance computing cluster at the University of Bern, Switzerland.

## Supporting information

**S1 Text. Supplementary appendix**. Further details about data sources, model, external validation, additional results and sensitivity analyses.

**S2 Text. TRIPOD checklist**. Reporting of model developing and validating according to the TRIPOD Checklist for Prediction Model Development.

